# Integrating Screening and Clinical Interviews: Advancing the Assessment of Exercise Addiction in Athletes

**DOI:** 10.1101/2025.10.28.25338951

**Authors:** Maria Geisler, Alexandra Arnold, Oliver Stoll, Birgitta Schiller, Eva Wimmer, Marco Herbsleb, Feliberto de la Cruz, Andy Schumann, Karl-Jürgen Bär

## Abstract

**Background and aims:** Questionnaire-based tools such as the Exercise Dependence Scale (EDS) are widely used to identify athletes exhibiting dependence-like features, but it remains unclear how these classifications relate to clinically relevant addictive exercise behaviour. This study combined quantitative screening with structured clinical interviews to (1) examine the proportion of athletes showing high EDS scores who meet ICD-11 criteria for a disorder due to addictive behaviour (6C5Y), (2) characterize psychological and motivational differences between athletes classified as exercise addicted versus non-addicted based on ICD-11 (6C5Y), and (3) identify the most discriminative predictors of this classification.

**Methods:** A total of 342 endurance athletes (159 female, 183 male) completed an online survey including the EDS, life satisfaction, and motivational factors. Athletes exceeding the EDS cut-off (>77; n = 63) and consenting to re-contact were invited to structured diagnostic interviews based on ICD-11 criteria for disorders due to addictive behaviors and additionally completed the SCL-90-R; 34 athletes participated in the interview phase (attrition rate: 46.0%).

**Results:** Of the 34 interviewed athletes, 24 (70.6 %, 95 % CI: 53.6–83.9 %) were classified as exercise addicted according to ICD-11, while 10 (29.4 %, 95 % CI: 16.1–46.4 %) were not classified as addicted. Compared to non-addicted athletes, those classified as addicted reported greater withdrawal-related symptoms, more comorbidities, lower life satisfaction, and higher weight-related exercise motivation, while training volume did not differ. In an exploratively LASSO model, withdrawal symptoms, past comorbidities, and interview-based anamnesis emerged as the strongest predictors of ICD-11 classification (AUC = 0.97).

**Conclusions:** High EDS scores often reflect exercise dependence features that may be normative in endurance athletes, whereas only a subset meet ICD-11 criteria for clinically relevant addictive exercise behaviour. These findings highlight the importance of withdrawal-related symptoms and comorbidities for differential assessment and support a multi-method approach to distinguish high involvement or dependence from true pathological addiction.

## 1. Introduction

Exercise addiction (EA) is a dysfunctional behavior characterized by excessive exercise, withdrawal symptoms, loss of control, and negative consequences such as injuries or social isolation (Heather A. Hausenblas & Danielle Symons Downs, 2002; Weinstein & Szabo, 2023). Despite these features, EA is not yet recognized as an independent diagnostic category in international classification systems such as the DSM-5 or ICD-11 (American Psychiatric Association, 2013). Nevertheless, it has attracted increasing scientific and clinical attention in recent years due to its potential overlap with established psychiatric conditions, including depression, anxiety, eating disorders, and obsessive-compulsive traits. Although the behavioral and psychological features of EA have been described in the literature, a number of critical gaps remain regarding its valid assessment and classification.

A central issue in the literature is the conceptual distinction between exercise dependence and exercise addiction, which is frequently unclear (Manfredi, 2022). Exercise dependence is conceptualized as “a craving for leisure time physical activity that results in uncontrollable excessive exercise behavior and that manifests in physiological and/or psychological symptoms” (Heather A Hausenblas & Danielle Symons Downs, 2002). The construct of exercise dependence, as operationalized in the Exercise Dependence Scale (EDS), is based on DSM-IV criteria for substance dependence, including tolerance, withdrawal, intention effect, lack of control, time investment, reduction of other activities, and continuation despite negative consequences. Importantly, in athletic populations, high scores on these dimensions might, in some cases, reflect adaptive or normative responses to intensive training, rather than reflecting clinically relevant addictive behavior (Allegre et al., 2006).

In contrast, exercise addiction refers to a clinically relevant disorder, in line with ICD-11 criteria for disorders due to addictive behaviours (6C5Y), which emphasizes impaired control, behavioral salience, persistence despite harm, and functional impairment.

To date, most research has relied on self-report questionnaires, such as (EDS) (Heather A. Hausenblas & Danielle Symons Downs, 2002; Müller et al., 2013) or Exercise Addiction Inventory (Griffiths et al., 2005). While these instruments have proven useful for screening, they are limited by their inability to distinguish between clinically relevant addiction and high, but non-pathological, levels of exercise or exercise (Müller et al., 2014; Weinstein & Szabo, 2023). This is particularly problematic in athletic populations, where high training volumes and sacrifices in other life domains may reflect professional dedication rather than psychopathology. As a result, high scores on the EDS may reflect dependence-like features of exercise rather than clinically relevant exercise addiction, and prevalence estimates based solely on questionnaire data may overstate the occurrence of addictive behavior in athletic populations. The only study to date directly comparing EDS-based classifications with structured interview outcomes was conducted by Müller et al. (2014). Their results indicated only fair to moderate agreement, suggesting that screening with the EDS may identify athletes with high involvement or dependence features, rather than true addiction. However, their investigation was limited to estimating concordance, without further exploring psychological or motivational differences between athletes classified as addicted versus non-addicted.

The present study aimed to replicate and extend the two-step assessment approach introduced by Müller et al. (2014). While their work demonstrated that EDS-based screening may identify athletes with high dependence features rather than clinically relevant exercise addiction, our study went further by exploring in detail how athletes classified as exercise addicted according to ICD-11 differ from those who screened “at risk” on the EDS but did not meet ICD-11 criteria. In doing so, we moved beyond prevalence estimation to examine psychological, motivational, and health-related characteristics that most strongly distinguish athletes meeting ICD-11 criteria from those who do not. Specifically, we aimed (1) to examine to what extent athletes classified as “at risk” by the EDS also demonstrated a clinically relevant pattern of addictive exercise behaviour according to ICD-11 criteria in structured interviews. Furthermore, we aimed (2) to examine differences between athletes with versus without a clinically classified dysfunctional exercise profile in biographical characteristics, life satisfaction, exercise dependence subscales, comorbid symptoms, and motivational factors, and (3) to determine which of these characteristics provide the greatest discriminative value, assuming that specific clinical features rather than general training behavior would be most informative.

## 2. Materials and Methods

### 2.1 Participants

Participants were recruited through social media platforms (e.g., Facebook groups for runners and triathletes), online sports forums, and other digital platforms, with a particular focus on addressing endurance athletes. Inclusion criteria for the online study were a minimum age of 16 years and engagement in at least four hours of sports per week. A total of 342 athletes completed the online questionnaire. To compensate participation, we raffled ten vouchers (50 € each) for a local running store.

For the interview study, all participants who scored above the established cut-off of EDS > 77 in the screening (N = 63) and who had provided consent to be re-contacted were invited. Ultimately, 34 athletes participated in the structured diagnostic interviews, see Fig. 1.

**Fig. 1.**
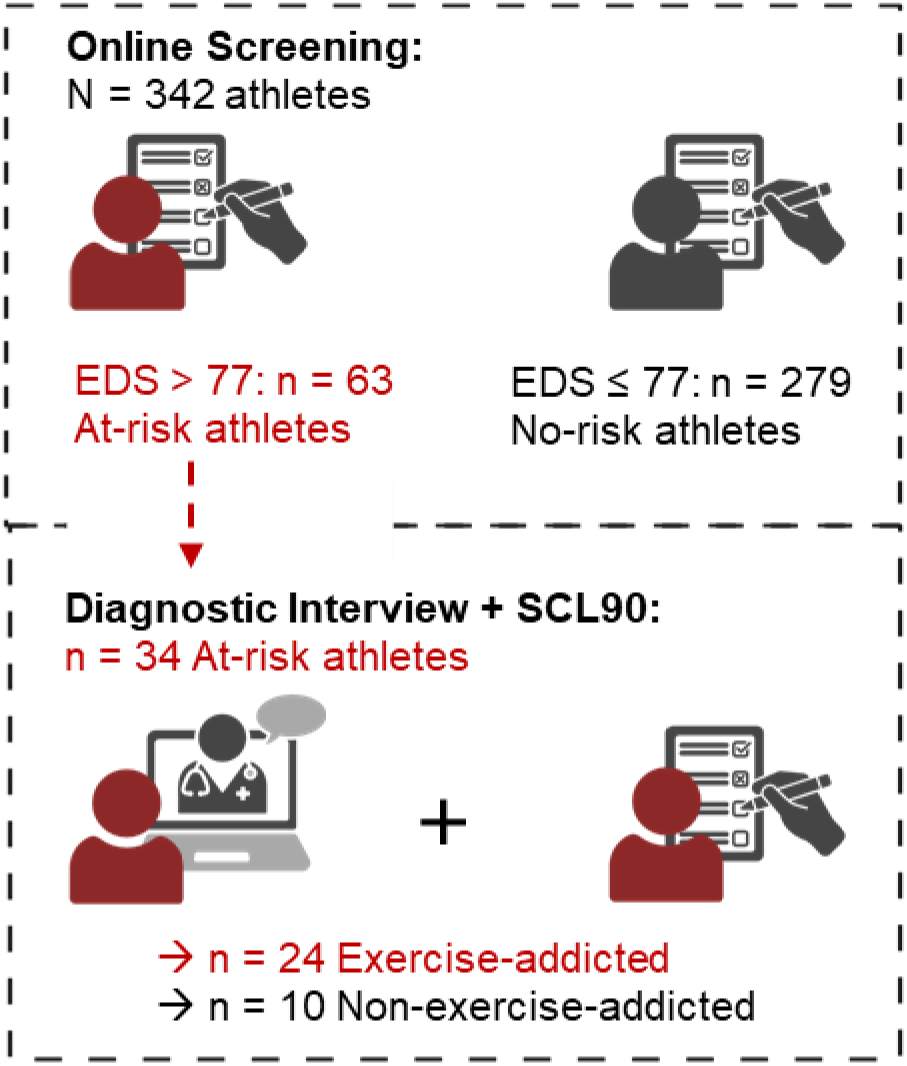
Study design. Following an initial online screening, athletes who scored above 77 on the EDS were invited to undergo a diagnostic clinical interview and complete the SCL-90. EDS = Exercise Dependence Scale; SCL-90 = Symptom Checklist-90.

All participants provided informed consent. The study was approved by the Ethics Committee of the University Hospital Jena (2024-3259_1-BO).

### 2.2 Study Design

The study consisted of two consecutive phases: an online screening and subsequent structured interviews.

*Online screening.* The web-based survey lasted approximately 10–15 minutes and included sociodemographic and training-related questions (e.g., weekly hours of endurance sport, number of workouts per week, years of practice), the Exercise Dependence Scale (EDS, including all subscales), life satisfaction, and sport-related motivational factors. Based on the EDS cut-off (>77), participants were categorized as at risk or no risk. Descriptive statistics of both groups are reported in Table 1.

**Table 1.** Characteristics of subjects based on online survey.

|  | At risk - athletes |  | No risk - athletes |  | *p-value* |
| --- | --- | --- | --- | --- | --- |
|  | *M* | *SD* | *M* | *SD* |  |
| Age (years) | 35.0 | 13.7 | 34.4 | 12.9 | 0.953 |
| BMI (kg/m<sup>2</sup>) | 22.8 | 2.6 | 22.4 | 2.3 | 0.608 |
| Exercise amount (h/week) | 9.7 | 4.2 | 9.6 | 5.4 | 0.359 |
| Exercise duration (years) | 15.9 | 13.0 | 13.0 | 9.2 | 0.284 |
| Exercise workouts (N/week) | 6.0 | 2.0 | 6.1 | 2.7 | 0.551 |
| EDS-Total Sum | 89.4 | 10.4 | 54.6 | 13.1 | **<0.001** |
| EDS-Withdrawal | 4.3 | 1.1 | 2.6 | 1.1 | **<0.001** |
| EDS-Continuation | 4.2 | 1.1 | 2.3 | 1.0 | **<0.001** |
| EDS-Tolerance | 4.2 | 0.9 | 2.6 | 1.0 | **<0.001** |
| EDS-Lack of Control | 4.2 | 1.0 | 2.3 | 1.0 | **<0.001** |
| EDS-Reduction in other activities | 3.7 | 0.8 | 2.3 | 1.0 | **<0.001** |
| EDS-Time | 5.2 | 0.7 | 3.8 | 1.1 | **<0.001** |
| EDS-Intention effect | 4.0 | 1.0 | 2.3 | 0.9 | **<0.001** |
| Life satisfaction | 5.6 | 1.1 | 5.3 | 1.2 | 0.201 |
| Motivation_health | 4.6 | 0.7 | 4.4 | 0.8 | 0.137 |
| Motivation_weight | 3.7 | 1.3 | 3.4 | 1.4 | 0.119 |
| Motivation_performance | 4.4 | 0.9 | 4.4 | 0.8 | 0.933 |
| Motivation_pleasure | 4.7 | 0.6 | 4.4 | 0.9 | **0.014** |
| Motivation_social | 3.3 | 1.4 | 3.1 | 1.4 | 0.252 |

Group-specific mean (M) and standard deviation (SD) of demographic variables in at-risk athletes and no-risk athletes. Group differences were assessed using independent-samples *t*-tests as all data were normally distributed. Significant *p*-values ( $< 0.05$ ) are highlighted in bold. BMI = Body Mass Index, EDS = Exercise-Dependence Scale.

*Structured interviews.* All athletes who (a) scored above the cut-off, and (b) consented to be contacted were invited for a follow-up clinical interview. A total of 34 interviews were conducted, each lasting 30–60 minutes. The interviews followed a semi-structured guide covering: (i) exercise addiction symptoms according to ICD-11 criteria (tolerance, withdrawal, impaired control, reduction of other activities, persistence despite injury/illness, secrecy); (ii) onset and triggers of problematic exercise behavior; (iii) potential comorbid conditions (eating disorders, obsessive–compulsive symptoms, substance use, others); and (iv) psychosocial background and psychiatric history. Before the interview, participants also completed the Symptom Checklist-90 (SCL-90) online.

### 2.3 Coding and Classification

Two independent raters (MG, AA) analyzed the interview transcripts using MAXQDA Analytics Pro (24.0.0). For the classification of clinically relevant addictive exercise behaviour, we applied the ICD-11 guidelines for “Disorders due to addictive behaviours” (6C5Y). Athletes were classified as showing a clinically relevant addictive exercise pattern if they fulfilled the essential ICD-11 features of impaired control over exercise behaviour, increasing priority given to exercise, and continuation despite negative consequences, persisting for at least 12 months and associated with significant distress or functional impairment.

For reasons of readability, the terms “exercise addicted” and “non-addicted” are used throughout the manuscript as descriptive shorthand to refer to athletes meeting versus not meeting these ICD-11 criteria. This terminology does not imply that EA constitutes a formally established diagnostic entity. Beyond this classification, additional codes were applied to capture relevant clinical features, including withdrawal symptoms, tolerance, current and past psychiatric comorbidities. Intercoder reliability was assessed based on the independent coding of both raters.

### 2.4 Statistical analyses

All statistical analyses were performed using R version 3.4.1 (Team, 2017). Significance levels were set to p ≤ 0.05.

In a first step, data from the online screening were analyzed descriptively, stratified by participants categorized as *at risk* (EDS > 77) versus *no risk*. Group differences between these two categories were examined using independent-samples *t*-tests or, when assumptions of normality were violated, Mann–Whitney *U*-tests.

In a second step, data from the structured clinical interviews were analyzed. Athletes classified as exercise addicted were compared with those classified as non-addicted (despite being categorized as *at risk* in the screening). Again, *t*-tests or Mann–Whitney *U*-tests were applied depending on distributional properties.

Finally, all variables that showed significant differences between addicted and non-addicted athletes in the interview-based classification were entered into a Least Absolute Shrinkage and Selection Operator (LASSO) logistic regression model. LASSO is a penalized regression technique that simultaneously performs variable selection and regularization by shrinking less relevant coefficients toward zero (Tibshirani, 2018). This approach is particularly suited for situations with many correlated predictors and relatively small sample sizes, as it reduces the risk of overfitting and multicollinearity.

We applied cross-validated LASSO logistic regression (10-fold cross-validation, *α* = 1) using the *glmnet* package in R. The penalty parameter λ was selected based on the “one-standard-error” rule (λ.1se), favoring a more parsimonious model. For variables retained in the final model (non-zero coefficients), we report standardized coefficients as well as odds ratios recalculated on the original scale of measurement to facilitate interpretability. Model performance was evaluated using classification accuracy and the area under the receiver operating characteristic curve (AUC).

Given the small sample size and the risk of model instability, we additionally conducted a stratified bootstrap stability analysis (1,000 resamples). In each bootstrap iteration, cases were resampled with replacement within outcome groups to ensure representation of both addicted and non-addicted athletes. For each resample, the LASSO model was re-estimated using the same cross-validation and penalty selection procedure. Selection probabilities for each predictor were calculated as the proportion of bootstrap samples in which the variable retained a non-zero coefficient.

## 3. Results

### 3.1 Screening-Based and Interview-Based Classification

A total of 342 athletes completed the online screening, of whom 63 (18.4%, CI: 0.248 – 0.229%) exceeded the EDS cut-off score (> 77) and were therefore classified as at-risk for exercise addiction (see Fig. 2). Descriptive characteristics of at risk-athletes and no-risk athletes are shown in Table 1.

**Figure 2.**
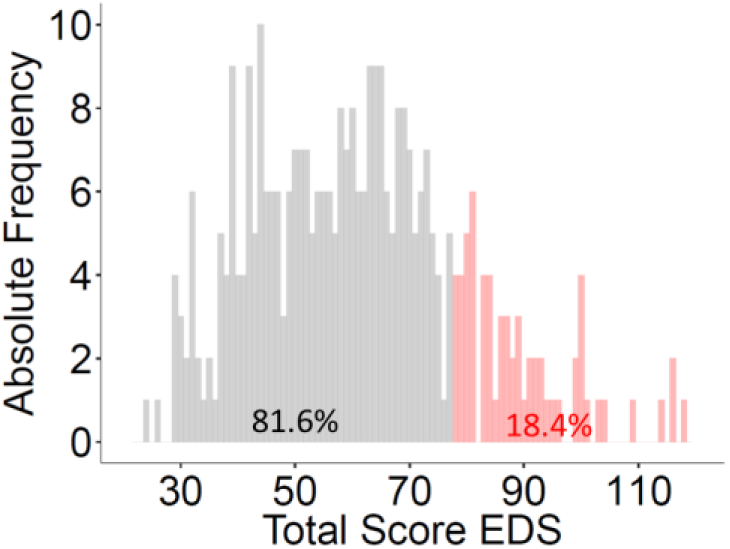
Screening-based classification of exercise addiction risk. Distribution of total scores on the Exercise Dependence Scale (EDS) across all participants (N = 342). Grey bars represent athletes classified as no risk (n = 279; 126 female, 153 male), and light red bars indicate those classified as at-risk (n = 63; 33 female, 30 male), based on the applied cut-off score of EDS > 77.

Of these at-risk classified athletes, 34 athletes agreed to participate in the follow-up clinical interview. Based on the ICD-11 criteria for “Disorders due to addictive behaviours,” 24 (70.6 %, 95 % CI: 53.6–83.9 %) were classified as exercise addicted, while 10 (29.4 %, 95 % CI: 16.1–46.4 %) were classified as non-addicted. Interrater reliability was 97% (agreement in 33 of 34 cases), confirming a high level of diagnostic consistency across both raters.

### 3.2 Group Differences: Exercise Addicted vs. Non-Addicted

All descriptive and inferential statistics comparing athletes classified as exercise addicted versus non-addicted are summarized in Table 2 and 3. Regarding demographic characteristics, the two groups did not differ in BMI (t(23.58) = −0.07, p =.947, d = −0.02 [CI =-0.72 – 0.76]), total exercise hours (W = 86.5, p =.208, r =.22 [CI = 0.02-0.5]), duration of athletic involvement (W = 97, p =.394, r =.15 [CI = 0.01-0.42]), or weekly number of workouts (W = 122, p =.954, r =.01 [CI = 0.01-0.39]). However, addicted athletes were significantly older than non-addicted athletes (W = 178.5, p =.028, r =.38 [CI = 0.1-0.61]). In terms of psychological and motivational variables, addicted athletes reported lower overall life satisfaction (W = 66, p =.037, r =.36 [CI = 0.06-0.63]), higher motivation related to weight control (W = 184.5, p =.012, r =.43 [CI = 0.17-0.69]), and lower performance-oriented motivation (W = 75, p =.029, r =.38 [CI = 0.23-0.53]). The two groups did not differ in social motivation (W = 82.5, p =.141, r =.26 [CI = 0.02-0.58]), fun-related motivation (W = 93, p =.297, r =.18 [CI = 0.01-0.49]), or health-related motivation (W = 110, p =.694, r =.07 [CI = 0.00-0.43]).

**Table 2.** Characteristics of interviewed at-risk athletes.

|  | Exercise Addicted |  | Exercise non-addicted |  | *p-value* |
| --- | --- | --- | --- | --- | --- |
|  | *M* | *SD* | *M* | *SD* |  |
| **Age (years)** | 34.3 | 12.4 | 22.6 | 4.6 | **0.028§** |
| BMI (kg/m<sup>2</sup>) | 21.3 | 2.8 | 21.4 | 2.0 | 0.953 |
| Exercise amount (h/week) | 12.4 | 7.7 | 12.4 | 2.6 | 0.208§ |
| Exercise duration (years) | 11.8 | 9.9 | 12.0 | 4.9 | 0.394§ |
| Exercise workouts (N/week) | 8.1 | 3.8 | 7.4 | 2.1 | 0.954§ |
| EDS-Total Sum | 91.7 | 9.5 | 84.9 | 6.4 | 0.053§ |
| EDS-Total Average | 4.4 | 0.5 | 4.0 | 0.3 | 0.053§ |
| **EDS-Withdrawal** | 4.7 | 0.8 | 3.7 | 1.2 | **0.036** |
| **EDS-Continuation** | 4.4 | 1.2 | 3.6 | 0.9 | **0.046** |
| EDS-Tolerance | 4.2 | 0.8 | 4.5 | 0.6 | 0.350 |
| EDS-Lack of Control | 4.4 | 0.9 | 3.7 | 1.0 | 0.107 |
| EDS-Reduction in other activities | 3.8 | 0.8 | 3.3 | 0.7 | 0.114 |
| EDS-Time | 5.1 | 0.7 | 5.4 | 0.5 | 0.287§ |
| EDS-Intention effect | 4.1 | 0.9 | 4.1 | 1.0 | 0.976 |
| **Life satisfaction** | 4.6 | 1.2 | 5.6 | 1.1 | **0.037§** |
| Motivation_health | 4.2 | 1.0 | 4.2 | 1.3 | 0.694§ |
| **Motivation_weight** | 3.9 | 1.5 | 2.7 | 0.9 | **0.012§** |
| **Motivation_performance** | 4.5 | 0.8 | 5.0 | 0 | **0.029§** |
| Motivation_pleasure | 3.7 | 1.1 | 4.1 | 1.0 | 0.297§ |
| Motivation_social | 2.2 | 1.5 | 3.0 | 1.3 | 0.141§ |
| **Current comorbidity** | 0.7 | 0.9 | 0 | 0 | **0.012§** |
| **Past comorbidity** | 1.1 | 0.9 | 0.1 | 0.3 | **<0.001§** |
| **Withdrawal sum** | 3.2 | 1.0 | 1.7 | 1.3 | **0.005§** |

Group-specific mean (M) and standard deviation (SD) of variables in addicted athletes and non-addicted athletes. Independent-samples *t*-tests or, when assumptions of normality were violated, Mann–Whitney *U*-tests (§) were used to test group differences. Significant *p*-values (< 0.05) are highlighted in bold. BMI = Body Mass Index, EDS = Exercise-Dependence Scale.

**Table 3.**
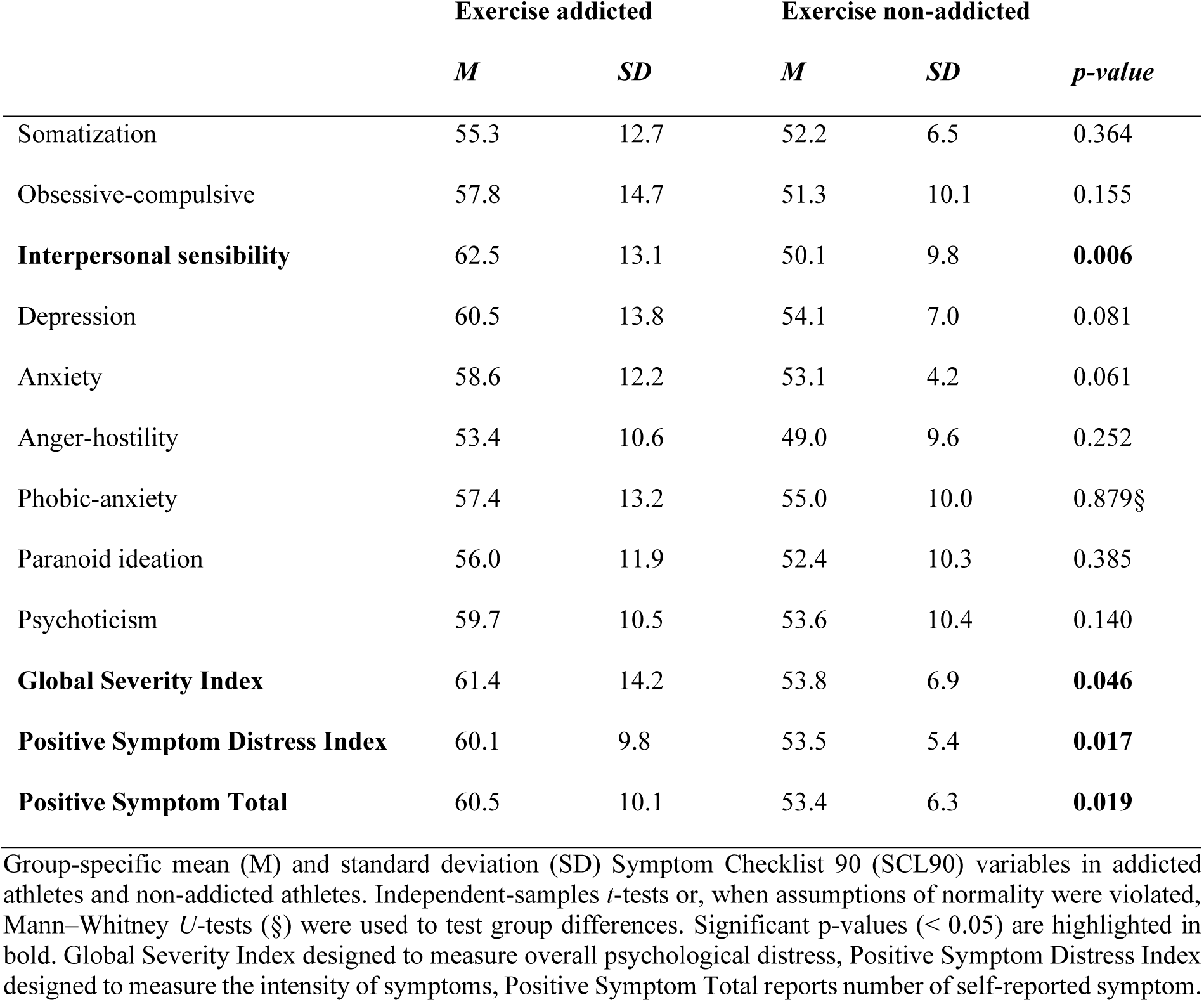
Symptom Checklist 90 characteristics of interviewed at-risk athletes.

Concerning exercise dependence, group differences were observed for the withdrawal (t(12.75) = 2.34, p =.036, d = 1.02 [CI = 0.24-1.80]) and continuation subscales (t(22.98) = 2.11, p =.046, d = 0.70 [CI = 0.07-1.45]), indicating higher scores among addicted athletes. No significant differences emerged for tolerance (t(22.55) = −0.95, p =.350, d = −0.32 [CI =-1.06-0.43]), lack of control (t(16.24) = 1.70, p =.107, d = 0.65 [CI =-0.11-1.40]), reduction in other activities (t(17.80) = 1.66, p =.114, d = 0.61 [CI =-0.15-1.36]), time (W = 92, p =.287, r =.19 [CI = 0.01-0.49]), intention effects (t(15.60) = −0.03, p =.976, d = −0.01 [CI =-0.75-0.73]), and the total EDS (W = 171.5, p =.053, r =.34 [CI = 0.05-0.60]), (see Figure 3). Regarding psychopathological symptoms assessed with the SCL-90, addicted athletes showed significantly higher scores on interpersonal sensitivity (t(22.43) = 3.01, p =.006, d = 1.01 [CI = 0.22-1.78]), on the Positive Symptom Total (PST) (t(26.55) = 2.49, p =.019, d = 0.78 [CI =-0.01-1.53]), the Global Severity Index (GSI) (t(30.98) = 2.08, p =.046, d = 0.60 [CI =-0.16-1.35]) and the Positive Symptom Distress Index (PSDI) (t(29.01) = 2.52, p =.017, d = 0.76 [CI =-0.01-1.51]) compared to non-addicted athletes.

**Figure 3.**
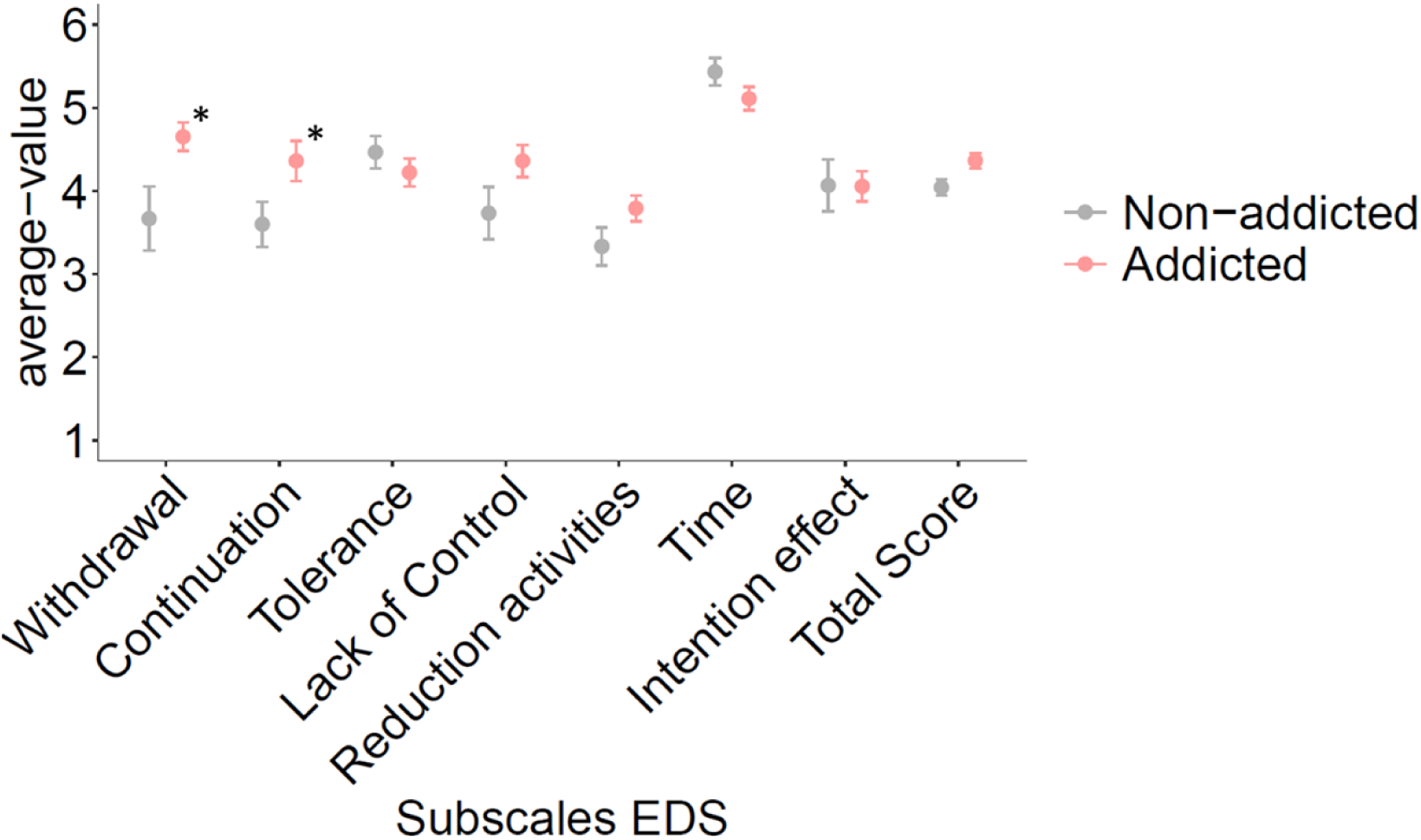
Exercise Dependence Scale (EDS) subscale scores (mean ± SE) for athletes classified as exercise-addicted (red) and non-addicted (grey). Significant group differences emerged for the total EDS and withdrawal subscale.

**Figure 4.**
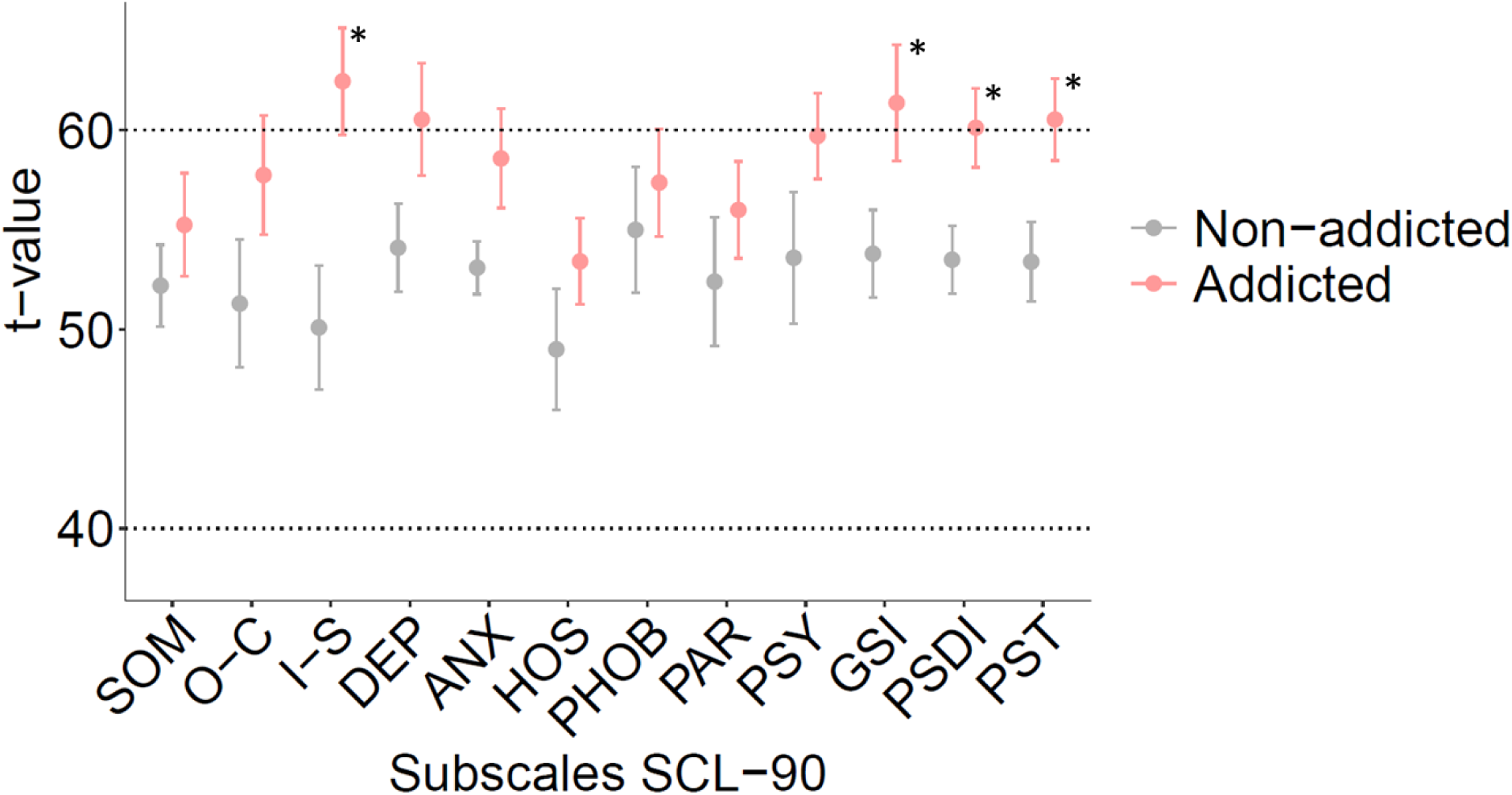
Mean T-scores (± SE) on the SCL-90 subscales for athletes classified as exercise addicted (red) and non-addicted (grey). Addicted athletes showed significantly higher scores on Interpersonal Sensitivity and the Positive Symptom Total (PST) compared to non-addicted athletes. T-scores are standardized values with a mean of 50 and a standard deviation of 10 (values ≥ 60 indicate clinically relevant symptom levels). SOM = Somatization; O-C = Obsessive–Compulsive; I-S = Interpersonal Sensitivity; DEP = Depression; ANX = Anxiety; HOS = Hostility; PHOB = Phobic Anxiety; PAR = Paranoid Ideation; PSY = Psychoticism; GSI = Global Severity Index; PSDI = Positive Symptom Distress Index; PST = Positive Symptom Total.

All other subscales showed no significant group differences, including somatization (t(30.33) = 0.92, p =.364, d = 0.27 [CI =-.47-1.01]), obsessive–compulsive symptoms (t(24.35) = 1.47, p =.155, d = 0.48 [CI =-0.28-1.22]), depression (t(30.48) = 1.80, p =.081, d = 0.53 [CI =-0.23 – 1.27]), anxiety (t(31.41) = 1.95, p =.061, d = 0.52 [CI =-0.23-1.26]), aggressiveness/hostility (t(18.52) = 1.18, p =.252, d = 0.43 [CI =-0.32-1.17]), phobic anxiety (W = 124.5, p =.879, r =.03 [CI =.01-.38]), paranoid ideation (t(19.54) = 0.89, p =.385, d = 0.31 [CI =-0.43-1.05]), and psychoticism (t(17.01) = 1.55, p =.140, d = 0.58 [CI =-0.17-1.33]).

Finally, analyses of interview-based data revealed that, beyond the ICD-11 criteria used for diagnostic classification, addicted athletes exhibited significantly more withdrawal-related symptoms (W = 192.5, p =.005, r =.48 [CI =.17-.71]), a higher frequency of current comorbidities (W = 175, p =.012, r =.43 [CI =.27-.60]), and more past comorbidities (W = 200.5, p =.001, r =.57 [CI =.34-.77]), compared to non-addicted athletes (see Figures 5 and 6).

**Figure 5.**
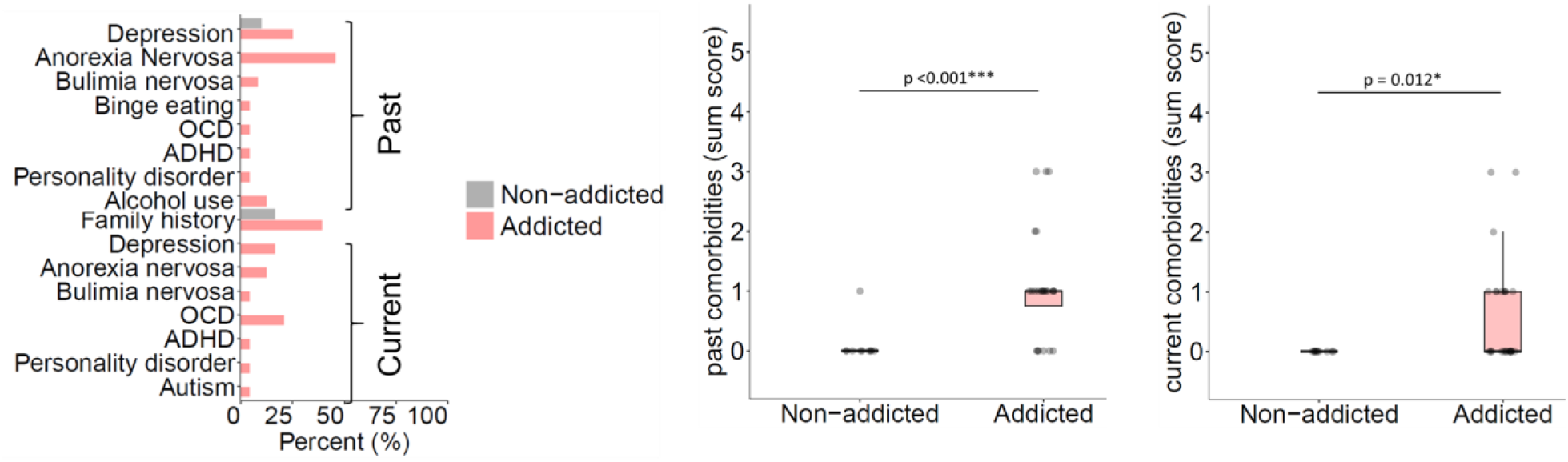
Past and current comorbidities among athletes classified as exercise-addicted non-addicted. Left: Percentage of athletes reporting specific comorbid diagnoses for addicted (red) and non-addicted (grey) groups. Center and right: Boxplots showing the summed number of past and current comorbidities, respectively. Athletes classified as exercise addicted reported significantly more past (p < 0.001) and current (p = 0.012) comorbidities compared to non-addicted athletes. OCD = Obsessive–Compulsive Disorder, ADHD = Attention-Deficit/Hyperactivity Disorder.

**Figure 6.**
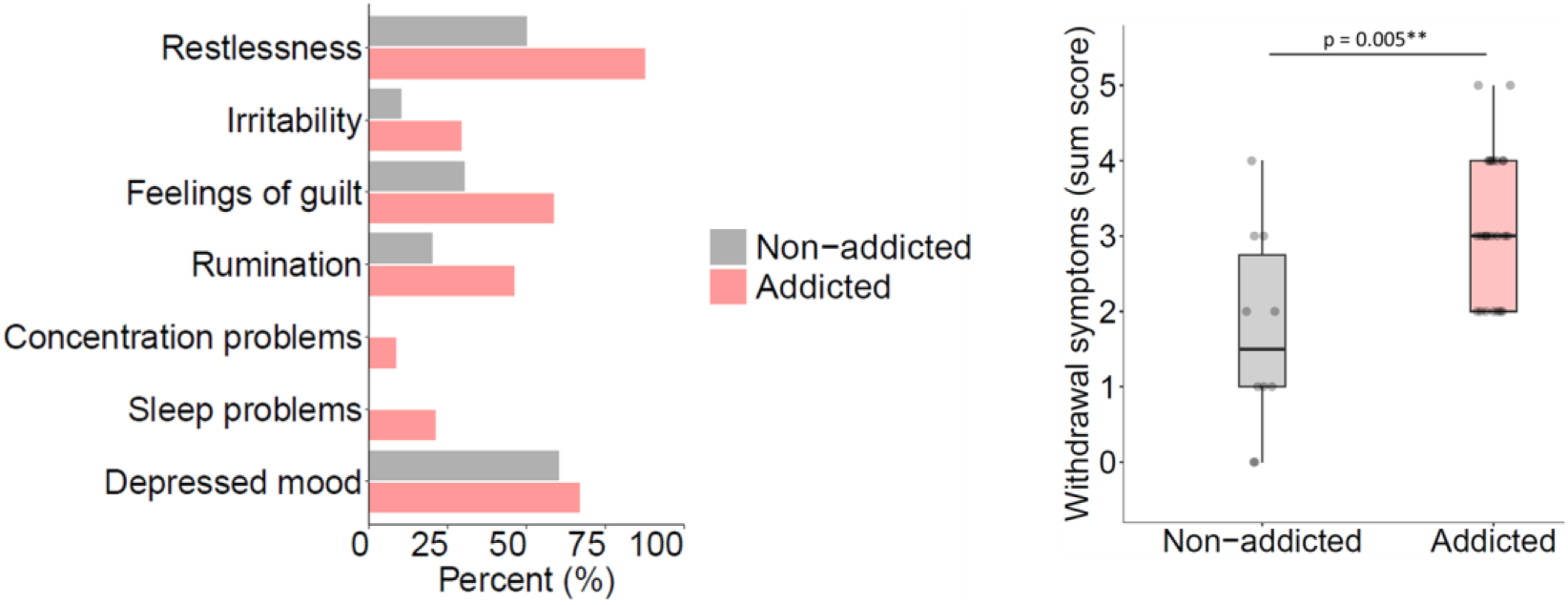
Withdrawal symptoms reported by athletes classified as exercise addicted and non-addicted. Left: Percentage of athletes reporting specific withdrawal symptoms. Right: Boxplots showing the total number of withdrawal symptoms reported by each group. Addicted athletes showed significantly more withdrawal symptoms compared to non-addicted athletes (p = 0.005).

### 3.3 Predictive modeling of exercise addiction

To identify the most informative predictors distinguishing between exercise-addicted and non-addicted athletes, a LASSO logistic regression was conducted including all variables that had shown significant group differences in previous analyses. The model was trained using 10-fold cross-validation to select the optimal degree of regularization (λ). The best-performing model was obtained at λ₁SE = 0.116, yielding a parsimonious set of predictors with non-zero coefficients. Two variables emerged as relevant predictors of classifying exercise addiction. These included the EDS withdrawal subscale ((β = 0.153, OR = 1.17), the interview-based withdrawal symptom sum score (β = 0.265, OR = 1.30) and the interview-based past comorbidity sum score (β = 0.279, OR = 1.32). The model achieved an overall classification accuracy of 94.1% and an area under the ROC curve (AUC) of 0.97, indicating excellent discrimination between addicted and non-addicted athletes. Given the small sample size (n = 34), a stratified bootstrap stability analysis (1,000 resamples) was performed to assess robustness. The bootstrap indicated that, in addition to the variables identified in the original model, age also frequently appeared as a selected predictor (selection probability = 63.1%). The interview-based withdrawal sum score had the highest selection probability (81.3%), indicating a robust and stable association with interview-based classification. The EDS withdrawal subscale (53.2%) and the interview-based anamnesis sum score (51.4%) showed moderate stability, suggesting potential relevance but requiring cautious interpretation.

## 4 Discussion

The present study combined questionnaire-based screening with structured clinical interviews to investigate EA in endurance athletes. Using a two-step design, we first identified athletes “at risk” according to the EDS and subsequently evaluated them against the ICD-11 guidelines for “Disorders due to addictive behaviours” (6C5Y) with a specific focus on addictive exercise behaviour. This approach yielded three main findings. First, only about two-thirds of athletes classified as “at risk” by the EDS were classified as exercise addicted in the structured interviews. Second, athletes classified as exercise addicted differed from their non-addicted counterparts mainly in psychological and motivational domains, rather than in training behaviour or anthropometric characteristics. Third, LASSO regression identified withdrawal symptoms and anamnestically reported comorbidities as the strongest predictors of ICD-11–based classification status.

### Diagnostic validity of questionnaire-based screening for exercise addiction

The present findings provide important insights into the validity and limitations of questionnaire-based screening approaches in the assessment of EA. Consistent with Müller et al. (2014), the proportion of athletes classified as “at risk” according to the EDS was highly comparable across studies (18.4% in the present sample vs. 17.2% in their study). This convergence suggests that the EDS reliably identifies a subgroup of athletes characterised by elevated dependence-related features across different samples and contexts.

However, the proportion of EDS-positive athletes who met interview-based criteria according to the ICD-11 framework differed substantially between studies. Whereas Müller et al. (2014) reported that only around 30% of individuals classified as “at risk” fulfilled interview-based criteria, this proportion was considerably higher in the present study (70.6%).

Several methodological differences may account for this discrepancy. Most notably, the present study used the ICD-11 framework, which provides more explicit behavioral and temporal criteria for addictive disorders than earlier diagnostic systems. Moreover, our sample consisted exclusively of endurance athletes, whereas Müller et al. (2014) examined a more heterogeneous group including fitness clients, sport students, recreational exercisers, and eating disorder patients. Differences in diagnostic framework, sample composition, and sport-specific norms may therefore substantially influence the proportion of EDS-positive individuals who meet interview-based criteria. Importantly, these findings should not be interpreted as evidence that the EDS is a flawed instrument. Rather, they underscore that the EDS primarily captures exercise dependence–related features, such as tolerance and withdrawal, which may be prevalent in endurance athletes without necessarily indicating clinically relevant addictive exercise behaviour (Heather A Hausenblas & Danielle Symons Downs, 2002). In this sense, questionnaire-based screening appears sensitive to high levels of exercise involvement and dependence-like patterns, but limited in its specificity for identifying clinically relevant addictive profiles when used in isolation.

### Psychological and clinical differences between EDS-positive athletes with and without clinically relevant addictive exercise behaviour

Beyond classification concordance, the present study provides a more differentiated picture by directly comparing athletes who screened positive on the EDS but differed with respect to interview-based ICD-11 criteria. Although both groups displayed comparable training volumes, duration of athletic involvement, and BMI, athletes classified as addicted were significantly older on average. This may indicate that dysfunctional exercise patterns develop gradually over time or become more salient with prolonged exposure to intensive endurance training. In line with previous research on exercise addiction (Lichtenstein et al., 2014), addicted athletes reported lower overall life satisfaction, a pattern also observed in other mental disorders (Meule & Voderholzer, 2020), suggesting reduced overall well-being despite high sport involvement. Importantly, given the cross-sectional design, these associations cannot be interpreted causally. Lower life satisfaction may represent a vulnerability factor, a consequence of dysfunctional exercise patterns, or a bidirectional process. The motivational profile further differentiated the two groups: exercise-addicted athletes exhibited stronger weight-related motivation and lower performance-oriented motivation. This shift from achievement-driven to weight-and control-oriented motives may reflect an underlying change from goal-directed to compulsive behavior, consistent with theories linking exercise addiction to affect regulation and body-related concerns rather than athletic performance (Costa et al., 2013; González-Hernández et al., 2023).

Interpreted through the lens of the Dualistic Model of Passion (Vallerand, 2015), this motivational shift further helps to distinguish between different qualitative forms of high exercise involvement. Athletes who screened positive on the EDS but did not meet ICD-11 criteria may be characterised by harmonious passion, marked by high commitment and identity relevance without substantial functional impairment. In contrast, athletes meeting ICD-11 criteria appear more consistent with obsessive passion, in which exercise engagement is driven by internal pressure, affect regulation, and difficulties disengaging despite negative consequences (Szabo & Demetrovics, 2022).

The pattern of EDS subscale differences supports this interpretation. Addicted athletes scored higher on the withdrawal and continuation subscales, indicating that negative reinforcement processes—exercising to avoid distress or negative affect—play a central role in maintaining the behavior. In contrast, tolerance, lack of control, and time investment did not differ significantly, possibly reflecting the structured and high-volume training routines that are normative in competitive sports rather than pathological.

Psychopathological differences were most pronounced in interpersonal sensitivity and global distress indices (PST, GSI, PSDI) on the SCL-90-R, pointing to heightened emotional vulnerability and social insecurity among addicted athletes. These findings are consistent with prior studies showing that perfectionism, interpersonal sensitivity, and self-esteem deficits contribute to compulsive exercise tendencies (Freimuth et al., 2011; Weinstein & Szabo, 2023). The absence of group differences in depression and anxiety may suggest that exercise continues to function as an emotion-regulation strategy, potentially masking affective symptoms in some athletes.

Interview-based data further underscore the clinical relevance of these findings. Athletes classified as addicted reported significantly more withdrawal symptoms and a higher frequency of both past and current comorbidities compared to non-addicted athletes. The relatively high odds ratios for past comorbidities in the LASSO model—together with interview-based withdrawal symptoms and anamnesis scores, which showed the highest selection probabilities in the bootstrap analysis—suggest that clinically relevant addictive exercise behaviour may be more likely to occur in the context of pre-existing psychological vulnerability. These results are consistent with conceptual models proposing that such behaviour could emerge as a secondary or compensatory phenomenon rather than as a purely sport-specific disorder (Weinstein & Szabo, 2023). However, given the small sample size and the exploratory nature of the LASSO analysis, these findings should be interpreted with caution and require replication in larger samples.

Taken together, the present findings highlight a crucial conceptual distinction between exercise dependence as a common and often normative feature of endurance sports and clinically relevant addictive exercise behaviour, which appears less prevalent and qualitatively distinct. From a motivational perspective, this distinction aligns with the differentiation between harmonious passion and obsessive passion (Szabo & Demetrovics, 2022; Vallerand, 2015). High dependence-related scores on the EDS may reflect adaptive commitment, harmonious passion, or professional dedication, whereas clinically relevant addictive exercise behaviour is characterised by obsessive engagement, withdrawal-related affective symptoms, comorbid psychopathology, maladaptive motivational patterns, and functional impairment.

### Clinical implications and diagnostic considerations

From a clinical perspective, these results underscore that screening tools such as the EDS are valuable for identifying athletes with elevated dependence-related features but should not be used as stand-alone indicators of addictive pathology (Weinstein & Szabo, 2023). The relatively high false-positive rate underscores the need for structured clinical interviews that incorporate e.g. ICD-11 criteria, including impaired control, increasing priority of exercise, and continuation despite harm. These features capture the qualitative aspects of addiction that go beyond frequency or intensity of training.

Second, the strong predictive role of withdrawal symptoms and comorbidity history suggests that assessment protocols should include systematic screening for psychiatric history and affective dysregulation. Clinicians working with athletes should be particularly attentive when exercise is used as a coping mechanism for anxiety, stress, or eating-related concerns, as these may signal the transition from high commitment to addictive behavior.

Third, the distinction between primary and secondary forms of exercise addiction deserves greater attention. Our findings that past psychopathology strongly predicts current EA align with conceptual models proposing that EA often emerges as a maladaptive coping strategy, particularly in individuals with prior affective or eating disorders. This perspective may have implications for treatment planning—suggesting that addressing underlying emotional regulation difficulties or body image concerns may be more effective than solely targeting exercise behavior itself.

Finally, the findings caution against the overpathologisation of dedicated athletes and argue for a multi-method assessment approach that integrates self-report screening, motivational analysis, and clinical interview data. Such an approach allows for greater diagnostic nuance and aligns with contemporary models emphasising dimensional, staged, and context-sensitive understandings of problematic exercise behaviour, including motivational frameworks such as the Dualistic Model of Passion (Szabo & Demetrovics, 2022).

### Limitations and Future Directions

Several limitations should be acknowledged. The number of clinically interviewed athletes was relatively small, which may limit statistical power and generalizability, especially regarding gender or sport-type differences. Although the interview demonstrated excellent interrater reliability (97%) and was grounded in ICD-11 criteria, it is not yet a standardized diagnostic instrument for exercise addiction. Future research should seek to refine and validate such interviews across athletic populations.

All assessments relied on self-report and retrospective recall, introducing potential biases due to social desirability or memory effects. Moreover, the cross-sectional design precludes causal conclusions—particularly concerning whether exercise addiction develops secondary to other psychopathology or vice versa. Longitudinal and experimental studies are needed to clarify these trajectories.

Although BMI was included as a descriptive measure, it is well-known that BMI has limited validity in athletic populations, particularly in endurance athletes (Bailey et al., 2025). BMI cannot distinguish between lean muscle mass and adiposity, which may obscure the true relationship between body composition, body image, and weight-related exercise motivations.

Finally, while the LASSO model identified a parsimonious set of predictors—withdrawal symptoms, comorbidity history, and interview-based anamnesis—external validation is essential to confirm their diagnostic value. Future research should also examine protective factors (e.g., coaching support, identity balance) and evaluate the applicability of emerging behavioral addiction frameworks, such as craving or functional impairment, to exercise-related disorders.

## Conclusion

The present study demonstrates that a considerable proportion of athletes who screen “at risk” on the EDS do not meet ICD-11 criteria for exercise addiction in clinical interviews, highlighting the limited diagnostic specificity of questionnaire-based screening. Addicted athletes differed from their non-addicted counterparts primarily in psychological and clinical characteristics—such as stronger withdrawal symptoms, lower life satisfaction, and more frequent comorbidities—rather than in training volume or duration. Moreover, past and current comorbidities emerged as the strongest predictors of addiction status, suggesting that exercise addiction may often develop in the context of, or as a maladaptive coping mechanism for, pre-existing psychological difficulties. These findings underscore the need to consider comorbid psychopathology and withdrawal-related features in the differential diagnosis of exercise addiction.

## Data Availability

All data produced in the present study are available upon reasonable request to the authors.

## Acknowledgements

We would like to thank Sissy Höbelt for her support in conducting the interviews.

## Data Availability Statement

The data of our study will be made available via a request to the Authors.

## Author contributions

MG, MH and K-JB designed the experiment. MG and K-JB collected the data. MG AA, MG, FD, AS, BS, EW analyzed the data. All authors discussed the results, revised the article critically for important intellectual content, and approved the final article.

